# Understanding Barriers and Facilitators of Primary Care Use Among Assertive Community Treatment Teams Via Qualitative Analysis of Clients and Clinicians

**DOI:** 10.1101/2023.12.05.23299368

**Authors:** Sophia Zhao, Walter Mathis

## Abstract

Individuals with severe mental illness and substance use disorders face complex barriers to achieving physical health. This study aims to explore the barriers and facilitators of primary care access among an Assertive Community Treatment (ACT) team. Semi-structured qualitative interviews were conducted with 14 clients and 7 clinicians from an ACT team at a community mental health center in Connecticut. Data analysis followed a grounded theory approach, with codes and themes emerging iteratively during the interview process. The study identified multifaceted barriers to accessing primary care, including economic challenges, homelessness, and the prioritization of mental health and substance use symptoms over healthcare. The conceptual framework consists of seven dominant themes: clients’ abilities, attitudes, experiences, knowledge, motivations, resources, and ACT-team level factors. This research provides valuable insights into the various barriers faced by ACT clients in accessing primary care. Improving primary care access for individuals with severe mental illness and substance use disorders is crucial for reducing health disparities in this vulnerable population.

## Introduction

People with serious mental illness and substance use disorders disproportionately experience comorbidities in cancer, cardiovascular disease, diabetes, obesity, and respiratory disease (1, 2, 3, 4). The increased morbidity and mortality due to medical causes seen in this population have been linked to factors including disparities in healthcare access, individual lifestyle choices, psychiatric symptoms, and side-effects of psychotropic medications (5).

Similar trends of poor physical health have been documented among individuals with serious mental illness served by Assertive Community Treatment (ACT) teams (6). ACT teams deliver care in the community, accommodating patients with serious mental illness and substance use disorders who have failed treatment in conventional outpatient settings (7). ACT teams are multidisciplinary, delivering case management, clinical treatment, and psychiatric rehabilitation services over long periods of time (7). Primary care serves as the entry point into the healthcare system and the continuing source of care for many patients with mental illness and substance use disorders (8). However, individuals with mental illness and substance use disorders have reported poor access to and quality of primary care (9, 10). Less is known about barriers to primary care for ACT clients, specifically. Understanding the distinct contexts underlying a population’s barriers to primary care can assist medical systems in effectively improving health outcomes (11, 12).

One study has collected information from ACT team members regarding barriers to physical health treatment among ACT clients (13). Other studies have assessed the integration of primary care-oriented interventions among ACT programs (14, 15). However, no studies have gathered input on barriers to primary care from ACT clients themselves. As engaging affected populations in research can help investigators address community priorities in a more effective, inclusive, and informed manner, further research is needed to better understand ACT clients’ perspectives.

Especially in ACT settings, both clients and clinicians play integral roles in the acceptability, effectiveness, and maintenance of primary care activities. In this study, we qualitatively explored barriers to primary care among ACT clients by conducting interviews with ACT team clients and clinicians at a community mental health center in New Haven, Connecticut.

## Methods

Interviews were undertaken with ACT team clients and clinicians at the Connecticut Mental Health Center in New Haven, Connecticut. Following consent to an interview, all participants received a ten-dollar gift card to Dunkin’ Donuts. All interviews were audio-recorded and transcribed either by hand by the researcher or a HIPAA-compliant transcription software. Each transcription was reviewed by the researcher for accuracy. This study was deemed exempt from IRB review by the Yale University Human Research Protection Program.

Client participants experienced severe mental illness, substance use disorders, frequent hospitalizations, and/or houselessness, and were recruited based on availability. Eligible client participants were sufficiently fluent in English to understand the consent form and participate in an interview. Client participants were either interviewed on-site at the Connecticut Mental Health Center (n = 12) or at their residence (n = 2). A majority of client participants were male (n = 11) and African-American (n = 8), followed by Hispanic (n = 3) and white (n = 3). All clinicians on the ACT team agreed to participate, with interviews conducted at the Connecticut Mental Health Center (n = 7). A majority of clinician participants were female (n = 6) and African-American (n = 3) as well as white (n = 3).

The semi-structured interview guide included several predetermined questions to explore themes related to barriers to primary care (see supplement). Follow-up questions were asked to probe deeper into issues brought forward by the participants.

De-identified transcripts were analyzed using a grounded theory approach. Data collection and analysis occurred simultaneously, allowing for the development and strengthening of themes (16). The first stage of our analysis involved analyzing the first few client transcripts using an open coding procedure where text reflecting barriers to primary care were organized into discrete codes. Then, relationships between these codes were identified and they were categorized under overarching themes (e.g., “Attitudes,” “Motivations,” “Logistics and Resources”). Codes and themes were iteratively refined throughout the interview process. After each revision, previously coded interviews were reviewed and adjusted as needed. Preliminary codes discovered during the analysis of client interviews were utilized as a starting point in the analysis of clinician interviews; the same process of iterative review and refinement was repeated. Where needed, the interview guide was amended to address emerging themes.

## Results

### Abilities

Both ACT team clients and clinicians discussed the impact of socioeconomic barriers on clients’ abilities to access primary care. Many clients relied on income support and were struggling to meet basic needs such as food and housing. (See Abilities Block A)

Participants further emphasized that symptoms stemming from severe mental illness or substance use disorders could prevent clients from prioritizing preventative care, let alone primary care. (See Abilities Block B)

Then, clinicians spoke about how clients’ physical health challenges were exacerbated from the outset due to the impact of psychotropic medications and social-structural circumstances including a lack of nutrition and the accessibility of unhealthy behaviors like smoking. (See Abilities Block C)

### Attitudes

Several beliefs and attitudes also emerge as barriers to primary care activities. Clients and clinicians alike described how a fear of both discovering health complications and medical intervention prevented clients’ use of primary care services. (See Attitudes Block A)

Another attitude that acted as a barrier to clients using primary care was clients believing that they were at low risk of serious physical illness. (See Attitudes Block B)

Clients additionally expressed discontent with the process of engaging with primary care facilitators, including the beliefs that it would be a waste of time, that physicians would invade their independence, and that ACT clinicians worked under ulterior motives. (See Attitudes Block C)

Alternatively, client participants also described attitudes that promoted the use of primary care, including being accustomed to primary care as a habit or tradition and seeing access to medical care at large as a human right and privilege. (See Attitudes Block D)

### Experiences

Both clients and clinician participants emphasized how past negative experiences in medical settings prevented ACT team clients from using primary care services. Namely, many ACT clients had previously experienced discrimination and traumatic events related to involuntary hospitalization, racism, and sexual trauma. (See Experiences Block A)

Client experiences with primary care were also profoundly influenced by their relationship with primary care providers that they encountered. Both clients and clinicians felt that clients were not “heard” or were misunderstood by their provider. (See Experiences Block B)

In many cases, this poor patient-provider relationship was rooted in a lack of clear, culturally-competent, and empathetic communication from the primary care doctor. (See Experiences Block C)

Client and clinician participants further described how the sheer number of medical services—for both physical and mental health—that ACT team clients received was overwhelming for them. Specifically, clients felt inundated with a high volume of appointments as well as discomforted by the nature of medical environments themselves. (See Experiences Block D)

Nevertheless, many clinicians described in detail how more positive experiences with primary care providers could improve clients’ use of primary care. (See Experiences Block E)

### Knowledge

Clients who did not regularly use primary care reported obtaining medical advice and guidance from alternative sources of knowledge, including from family and popular media. (See Knowledge Block A)

Clinicians offered a different perspective to clients’ knowledge of primary care, reporting that clients often lacked an understanding of the benefits and importance of primary care. Specifically, clinicians described how clients’ challenges to conceptualize their health outcomes played a role in implementing primary care activities. (See Knowledge Block B)

Moreover, clients were perceived to lack knowledge of how primary care could function as a source of health maintenance rather than negative diagnostic news. (See Knowledge Block C)

Clinicians additionally discussed how there is often little perception of the connection between mental and physical health. They emphasized the need for medical systems to place greater importance on a holistic view of healthcare for patients. (See Knowledge Block D)

### Motivations

Client and clinician participants largely differed in their perceptions of clients’ motivations to use primary care. Many clients spoke about their willingness to engage with primary care due to factors such as building positive relationships with their primary care provider. (See Motivations Block A)

A meaningful aspect of these strong relationships was the willingness of primary care providers to assist patients in a range of needs, from care coordination to education. Many clients expressed confidence in primary care providers’ abilities. (Motivations Block B)

Clients also expressed a sense of personal responsibility to maintain health. (Motivations Block C)

Another motivating factor was the opportunity to resolve the threat of disease, a point which clinicians raised as well. (Motivations Block D)

However, clinicians reported that clients often lacked an interest in and the motivation to use primary care activities. (Motivations Block E)

Clinicians described how clients came to rely on external motivation, such as food or money, as a precursor to engage with primary care. (Motivations Block F)

Furthermore, clinicians discussed how clients were less motivated to engage with primary care services due to a lack of support from family members. (Motivations Block G)

Several clinicians also emphasized the importance of fostering ACT team clients’ self-confidence, especially with regard to perceiving primary care as a non-threat. (Motivations Block H)

### Resources

Both clients and clinicians spoke extensively about how a lack of specific resources, such as cell phones and transportation, could limit clients’ ability to access primary care. (Resources Block A)

Then, clinicians highlighted how the limited nature of certain insurance plans that clients were under prevented them from utilizing a full range of primary care-related services. (Resources Block B)

### ACT Team-Level Factors

Interviews with clinicians indicated that clients’ ability to access primary care services was strongly influenced by the capacity of the ACT team to provide them assistance and direction. However, several clinicians indicated that a lack of coordination or inconsistencies in electronic record-keeping presented a barrier to performing this task. (ACT Team-Level Factors Block A)

Despite this technical setback, clinicians expressed confidence in their ability to assist clients in navigating primary care, including accompanying them during appointments and explaining medical concepts to them. (ACT Team-Level Factors Block B)

Another strength cited by clinicians was their ability to normalize primary care. This included not only regularly reminding clients to make medical appointments, but also having available a nurse who could provide medical tests including bloodwork. (ACT Team-Level Factors Block C)

Nevertheless, some clinicians highlighted the need for the ACT team to improve on its ability to normalize primary care among clients. (ACT Team-Level Factors Block D)

Finally, many clinicians stressed the importance of building rapport with clients and respecting their beliefs and values as crucial components of buy-in into medical care. (ACT Team-Level Factors Block E)

## Discussion

In this study, we identified barriers and facilitators of primary care service use among ACT team clients, elucidating contributors to the trend of worse physical health among individuals with severe mental illness and substance use disorders treated by ACT teams.

**Abilities:** Economic challenges, homelessness, and struggles to meet basic needs hindered clients’ ability to prioritize primary care. Symptoms stemming from mental illness or substance use disorders often took precedence over healthcare.
**Attitudes:** Clients expressed various attitudes that acted as barriers, including fear of discovering health complications, concerns about medical interventions, and mistrust of healthcare providers. Conversely, some clients recognized primary care as a natural and necessary aspect of their overall health.
**Experiences:** Negative past experiences in medical settings, such as discrimination and traumatic events, deterred clients from seeking primary care. Poor patient-provider relationships, often characterized by a lack of empathy and effective communication, further discouraged clients from primary care services.
**Knowledge:** Clients used alternative sources of knowledge, including family and popular media, for medical advice, often lacking an understanding of the benefits of primary care. Additionally, clients sometimes did not connect their mental and physical health.
**Motivations:** Motivations to engage with primary care varied among clients. Some were willing to engage due to positive relationships with providers and a desire to prevent disease. Others lacked motivation and relied on external incentives.
**Resources:** Limited resources, such as financial coverage by insurance, hindered clients’ access to high-quality primary care. The availability of cell phones and transportation were particularly crucial for appointment attendance.
**ACT Team-Level Factors:** The capability of ACT teams to provide assistance and coordination played a significant role in clients’ ability to access primary care. Building rapport, normalizing primary care, and respecting clients’ beliefs and values were identified as important factors in facilitating primary care engagement for ACT clients.

Altogether, these findings shed light on the multidimensional barriers that individuals with severe mental illness and substance use disorders face in accessing primary care. The economic, social, and psychological challenges highlighted in this study underscore the need for tailored interventions to improve healthcare access for this particularly vulnerable population.

These results align with previous research that has identified barriers to primary care for individuals with serious mental illness and substance use disorders. For instance, one group identified that clients, providers, and healthcare systems all played an integral role in facilitating primary care use among individuals with mental illness and substance use disorders (8). Furthermore, our findings corroborate the impact of negative past experiences on healthcare-seeking behavior. ACT clients’ reports of discrimination and traumatic events within medical settings align with previous research that has documented the stigmatization of individuals with mental illness and their subsequent reluctance to engage with healthcare (17, 18).

Similarly, the significance of the patient-provider relationship echoes literature that emphasizes the pivotal influence of empathetic and supportive providers on people with mental illness and substance use disorders (19).

Our study reaffirms these results and adds granularity to existing research by providing insight into a specific cohort of ACT clients. While previous research has often relied solely on the views of healthcare providers (20), our investigation directly engages with ACT clients who receive care, allowing for a more comprehensive understanding of their specific challenges and needs. Namely, this research uncovers the role of ACT clients’ experiences, motivations, and knowledge as underexplored barriers to primary care use. Moreover, our study underscores the significant function of the ACT team itself in hindering or facilitating primary care access. This focus on the organizational dynamics related to ACT teams adds a unique perspective to the existing literature, highlighting the importance of cohesive and supportive healthcare teams.

Despite these strengths, it is important to acknowledge that this qualitative study, focusing on a single ACT team from in New Haven, Connecticut, may not generalize to the broader population of individuals with severe mental illness or substance use disorders. As such, our findings may not capture the full spectrum of barriers faced by individuals with similar conditions in different treatment contexts. Furthermore, participants were recruited based on availability and willingness to participate, which may have excluded individuals with more severe barriers to primary care access. Finally, the qualitative nature of this study exposes it to potential bias. The personal perspectives of the researcher conducting the interviews may have influenced the data collection and interpretation processes. Steps were taken to minimize this effect through training and regular debriefing among the research team.

The findings of this study have several implications for healthcare practice. First, addressing practical barriers is vital. ACT teams can collaborate with community organizations to provide reliable transportation options for clients (21). Ensuring that clients have access to cell phones can also enhance communication and appointment scheduling (22). Then, our research suggests that further provider education is needed.

Many ACT clients felt that primary care providers lacked understanding of mental illness and substance use disorders. Training for healthcare providers should emphasize de-stigmatization of such health conditions and trauma-informed care (23, 24). Training programs should equip providers with the skills to engage with patients effectively and respectfully. Next, interventions should focus on providing ACT clients with comprehensive education about the importance of primary care and managing physical health conditions. Previous research has suggested that such educational initiatives should be highly-tailored, avoid overloading clients with information, and offer ample time for discussion and reflection (25). Lastly, ACT teams should establish concrete mechanisms for soliciting client feedback on their primary care experiences. Such information can be used to continuously identify areas for improvement in care coordination, communication, and service delivery. ACT teams must uphold its goals of patient-centered care, ensuring services and systems are meeting clients’ needs.

Further research is required to determine the effectiveness of any such interventions. Additionally, comparing the barriers and facilitators of primary care among ACT teams across different geographical locations may highlight best practices.

## Conclusions

Our study contributes valuable insights into the barriers faced by ACT clients in accessing primary care. These findings highlight the complex interplay of psychological, provider-level, and socioeconomic factors that influence primary care utilization in a population of ACT team clients. Addressing these barriers requires a multi-faceted approach that includes practical interventions and healthcare system reform. Ultimately, improving primary care access for individuals with serious mental illness and substance use disorders is essential for reducing health disparities and enhancing overall well-being in this vulnerable population.

**Table 1.** Client and provider quotes arranged by theme and block.

| Theme | Block | Quotes |
| --- | --- | --- |
| Abilities | A | “I haven’t gotten around to it. Just looking for a job.” (Client)<br>“Some of them don't have houses or [...] they’re out panhandling [...] they’re roughing it out. So to deal with something [like primary care] that they really don’t think is a priority just ends up being to them, I think, a nuisance.” (Provider)<br>“Some of our folks have conservators [...] [it] might be a struggle just getting [...] entitlements turned on and making sure that nothing happens to get it shut off.” (Provider) |
|  | B | “I don’t think I could [go to the dentist] by myself mentally.” (Client)<br>“It's kind of difficult to say [how I maintain my physical health] now because I have mental health problems now so I can't really do what I used to.” (Client)<br>“I can only imagine trying to keep myself organized enough with all of this noise in my head [...] that some other things become not a priority at all [...] Like I just gotta get through the day, who cares about brushing my teeth. I just gotta get through the day and function to some extent. Getting my teeth cleaned, brushing my teeth, or even showering, all that [...] is extra exhausting I would imagine.” (Provider)<br>“The substance abuse [...] if they’re using, then [...] they have very little insight [into improving their health]. If they’re not doing well psychiatrically and they’re distracted by internal stimuli, that’s going to be tough.” (Provider) |
|  | C | “A lot of our clients’ medication certainly does dry their mouths out, so it’s a greater risk of them developing [...] gum disease.” (Provider)<br>“[Psychotropic medications] cause weight gain [...] And people with mental illness tend to die earlier because of [...] those diseases, hypertension, diabetes.” (Provider)<br>“With poverty it’s hard to eat right. You know if you have a fixed income, you can’t buy all the fresh fruits and vegetables, you buy canned [food] [...] Some of our folks don’t really know how to cook so they buy easily-prepared stuff.” (Provider) |
|  |  | “A lot of the corner stores [take] food stamps to give people cigarettes.” (Provider) |
| **Attitudes** | A | “Maybe [I will get a medical screening] when I [...] want to make the choice. Right now, [...] I don’t think I want to like, complicate like, any situations.” (Client)<br>“What am I going to do to address [the chronic pain in my left leg?] I don't know, I don't want to go into that, I don't want to go into that [X-ray] machine. I don't want to go into there [...] It’s just strange. Strange. I don’t like that.” (Client)<br>“Our folks are [...] petrified of the dentist. They [...] just fear like the tools and stuff. It's [...] just being in the vicinity of their dentist [which] is anxiety-provoking.” (Provider)<br>“Some people don't want to know [about their diagnosis] [...] so they don't go to the doctor [...] A lot of people would rather find out later and deal with the consequences in that short amount of time. When it's too late.” (Provider) |
|  | B | “I’m not worried about [cancer or heart disease]. I’m not worried about anything like that. I’m one hundred percent healthy.” (Client)<br>“I just feel like I have [...] a couple more years left to be in good shape. I have a couple more years [...] before I have to worry about health problems.” (Client) |
|  | C | “I hate going to the doctor because I feel like it’s a waste of time.” (Client)<br>“[Doctors] [...] come in with other patients they [are] in control over. I don’t [...] [want] to be [under] control I just want to be in and out [after] ten minutes.” (Client)<br>“[ACT clinicians and providers], they [are] worse than anybody else, they [are] only here for the paycheck.” (Client) |
|  | D | “[Going to primary care] just feels natural now. I’ve known a primary doctor since I’ve been growing up so it’s just nothing new, nothing different.” (Client)<br>“Exams are very needed in America [...] Some people leave that to the end. Some go through it at the beginning. I think it’s a good thing.” (Client)<br>“Some people don't have medical care, so I appreciate it [...] Definitely feel grateful. Thankful and grateful [...] Everybody should have medical care. Everybody [doesn’t] have medical care, though.” (Client) |
| **Experiences** | A | “It was crazy they took me in there [and] strapped me down to the bed, beat me up, broke my ribs, collapsed lungs, broken hand [...] it was just crazy.” (Client)<br>“I wish [the hospital] just had a camera so you could see the difference [between] Black patient and white patient [...] you [would] see the difference [in] how they treat a white person and how they treat a minority.” (Client)<br>“If I’m a female you should bring a female to search me because it’s a right everywhere you go. At one point [...] [the hospital] wouldn’t bring a female so the guy searched me the wrong way.” (Client)<br>“I think it's a lot of paranoia [...] [how] if they see the doctor, they’re going to be told [...] they’re going to be put in a hospital against their will.” (Provider) |
|  | B | “Every time I try to get my point across [...] they want me to answer their questions but they don’t want to answer my questions.” (Client)<br>“I think it's just a lack of knowledge. [Providers] just think somebody’s being rude and they don’t have time [...] Instead of realizing that, you know, it took a lot to get this patient here because they’ve got other stuff going on.” (Provider)<br>“If [clients are] coming in with somatic symptoms, then they’re told [...] it’s psychosomatic. It’s not real. So [providers are] attributing it to their mental illness rather than, you know, there is this pain here.” (Provider) |
|  | C | “When you ask [providers] what’s wrong with you, they don’t break it down for you. I think that should be [...] especially important for people like me.” (Client)<br>“Sometimes in the past when I was going with patients to appointments, [providers] would talk to me instead of the patient.” (Provider)<br>“But [some doctors are] talking like a two-year-old. And I think for some [clients], you know, they’re grown men and it’s like, treat me like an adult. You know, I have a mental health [condition] but I'm still an adult [...] I’ve seen it many times when [doctors] talk down to [clients].” (Provider) |
|  | D | “Just things get overwhelming, you know? [...] I just go to the doctor too much. Or, I haven’t been to the doctor too much but I know I come here [to the Connecticut Mental Health Center] every Tuesday so that’s a lot [...] to bear sometimes.” (Client) |
|  |  | “Some of our folks [...] get to a point where they’re overloaded with supports and they’re just like just a hundred percent done, they don’t want to do anymore, they don’t want to discuss anymore issues” (Provider)<br>“It’s scary in [the hospital] [...] It’s like people [are] coming out anywhere, popping up, getting searched and [being put] in a room. Some people cry, some people [are] using phones, some people [are] watching TV, some people [are] eating.” (Client) |
|  | E | “I’ve had one primary care physician with a client who was very involved [...] he would call me and say [...] so-and-so didn’t show up for her appointment. Can you encourage her to come in? Can you bring her? [...] He would take the extra step.” (Provider)<br>“But I find the doctors that we bring most of our clients to [...] they’re respectful. They [...] tend to the needs of the clients. They ask questions. They try to have the clients follow through with the recommendations.” (Provider) |
| **Knowledge** | A | “My favorite doctor, Dr. Sebi [a popular healer and herbalist in the media], says that as long as you got mucus in [...] your body parts, if you got mucus in there, then you’re safe.” (Client)<br>“[If I ever got any serious illnesses] I’d just pray, take a lot of herbs, and diet change. And as long as I can walk and clean myself then I’m okay.” (Client)<br>“I [go] and ask my sister about [my problems] because she knows so much, she’s a nurse [...] always ask her about stuff.” (Client) |
|  | B | “I think the biggest barrier is [clients] not having insight into their medical condition. They don’t feel like it’s a need. It’s just a nuisance [...] So to just schedule another appointment with another provider, they don’t feel like it’s a need to begin with.” (Provider)<br>“It’s like if [clients] get a diagnosis, they’re not sure what it is, they don’t know how it’s going to affect them, so they just avoid it.” (Provider) |
|  | C | “Because [clients are] not going [to appointments] when they’re healthy [...] they’re always going to get bad news at the doctor, they don’t know you can go and just be okay, just to make sure you’re healthy.” (Provider) |
|  | D | “I’ve seen the big connection between your physical and mental health, and I don’t think that our clients get that [...] I really don’t think they have any idea [...] that they even need to go for physical health.” (Provider) |
|  |  | “Before I started working with the ACT team, I was working with the Behavioral Health Home. The whole model is treating the whole person, not just their mental health, but everything [...] So I'm hoping that there is a slight shift in that direction.” (Provider) |
| **Motivations** | A | “[I’m most satisfied with] the socializing thing, the one-on-one [...] how I’m doing, what progress I’m making.” (Client)<br>“I can discuss anything with her [my primary care provider], that’s why I like her. I think she would make a good therapist also.” (Client) |
|  | B | “If it wasn’t for [my primary care provider] I would have to track different departments and different types of doctors [...] It would be really difficult.” (Client)<br>“Anytime I went through a problem they would always do whatever they could [...] I never had a doctor who just didn't know what they were doing.” (Client)<br>“We talk and she [my primary care provider] helps me out and she tells me what I should do, like drink water, to keep hydrated, stuff like that.” (Client)<br>“[My primary care clinic] ha[s] information on eating healthy and stuff like that [...] better understanding about the foods I eat, the meds I take.” (Client) |
|  | C | “[If I got any health problems], I’d go to the primary care doctor [...] I’m not trying to fall back [...] to get back down to the negative side of [...] being sick.” (Client)<br>“[Going to the primary care doctor has] been really very good for me [...] It gives me a chance to find out how healthy I am, what could be wrong with me, what's not wrong with me, what could go wrong with me.” (Client)<br>“I think for everybody, they should [...] focus on themselves [as] being the most important person for their health.” (Client) |
|  | D | “I have to [get screenings] again [...] I want to know where it is right now because I haven’t been taking my medication for [...] more than a year.” (Client)<br>“I just don’t want the rest of my teeth to fall out [...] So I was hoping to get whatever they [the dentist does].” (Client)<br>“When they go [to their appointments], they wait until something’s wrong, and that’s why they re-associate with the doctor.” (Provider) |
|  | E | “What’s the most challenging? Just getting them to buy in. Sometimes you gotta go round and round [...] several times before you put it on the shelf.” (Provider)<br>“I also think a lot of them just don’t [...] even care about their physical health.” (Provider) |
|  | F | “What [the ACT team] tr[ies] to do is [use] gift cards, or going out to lunch [...] That seems to be what works the best for our clients in the beginning, just to engage them with the doctor.” (Provider) |
|  | G | “We have clients, both male and female, where their significant other might discourage them [...] from keeping their medical appointments or discourage them from keeping the medical follow-ups.” (Provider)<br>“We could do things for [unnamed ACT client], but we have to go to the [primary caregiver] and she’s not on the same page as us. So a lot of times we have to sit back. And by the time she comes to us, things are so bad, you know, [he’s] losing all his teeth [which] shouldn’t have happened if we were working [on] it two years ago.” (Provider) |
|  | H | “Sometimes once they get in the door and they see it’s not as scary as they had imagined or remembered [...] then you can get them to buy in more.” (Provider) |
| **Resources** | A | “I get a ride [to the clinic] from where I’m staying at. Staff usually bring me. Sometimes I take the bus. Depends on whether I have a bus card or not.” (Client)<br>“If I have any questions I have a number I can call if I need whatever information [...] If I don’t call in they’ll call me to schedule [an] appointment for me to make sure that everything is working well, what I’m taking.” (Client)<br>“[I haven’t kept up with appointments because] I haven’t had a phone, I haven’t had transportation.” (Client)<br>“The bus passes haven’t come for over a month for our folks and the buses are expensive [...] if a person can’t take the bus [a company] would do the driving transportation for them but sometimes they would have to wait, I heard one person had to wait for up to two hours. Then they start missing their appointments.” (Provider) |
|  | B | “So dental, that seems to be a big issue, finding good dental that accepts state insurance [...] or some of the specialties will be like, there are so many people on the waitlist to get maybe a neurological assessment that they’re waiting [...] months to get care or get assessed.” (Provider) |
|  |  | “Instead of fixing the tooth, the doctor will pull out all their teeth and give them dentures because it’s cheaper for them to do that.” (Provider) |
| **ACT Team-Level Factors** | A | “Not everybody has access [to electronic medical records] [...] I think it makes a huge difference [...] that way we know what the last doctor said to them, what they were looking at, and what they need from us to help that person meet their needs [...] we [instead] have half paper chart half electronic chart [...] that set back everything [...] us being efficient in trying to help people get their medical needs met.” (Provider) |
|  | B | “We’ll go inside with them [...] and then we’ll explain to the clients [...] what [...] the follow up recommendations are.” (Provider) |
|  | C | “I [...] do a very general assessment, check vitals, see [...] when was the last time they saw their primary care? Do they have primary care? [...] [I am] making it clear to clients [...] I’m here for this. [...] So let’s spend five, ten, fifteen minutes talking about, you know, let’s check your blood pressure. Talk about your dental stuff and your nutrition and your exercise level and your smoking.” (Provider) |
|  | D | “[Clients] might not want to do it [go to primary care], but I think that we should promote it more so that they at least have the option in their head.” (Provider)<br>“Maybe have more groups that engage in physical activities [...] maybe the team itself can do more [...] or having a health group.” (Provider) |
|  | E | “It’s [...] being just supportive, understanding people, talking to them about their fears [...] and we get it, we know your experiences from the past.” (Provider)<br>“Okay, so if, like, the person doesn't want to, for instance, [a] colonoscopy [...] then I can say [...] Maybe you can give a stool sample and we can do it that way. So that’s respecting that person’s decision to not want to get one. Still educating them and still trying to find a way that they can still receive some sort of care.” (Provider)<br>“I had a client comment to me [...] you’re the fifth worker I’ve had on the ACT team, so they notice stuff for sure [...] Rapport with a person is so important, and it makes a difference using providers that they are familiar with.” (Provider)<br>“Every two years or every year somebody [leaves] and we get a new fellow every year and then the clinicians leave and it’s hard. By the time you form that relationship, they’re leaving and then you start over. So they don’t have that trust.” (Provider) |

## Supporting information

Supplement Questions

## Data Availability

All data produced in the present study that do not contain protected health information are available upon reasonable request to the authors

