## Supplement Questions for "Understanding Barriers and Facilitators of Primary Care Use Among Assertive Community Treatment Teams Via Qualitative Analysis of Clients and Clinicians"

Questions asked to clients:

- How do you define primary medical care?
- Do you have a primary care doctor?
- Have you ever found it difficult to get primary medical care?
- How do you feel about going to your primary care doctor?
- What has your experience been like going to the primary care doctor?
- Can you tell me about the last time you went to the primary care doctor?
- Can you tell me about what you did the last time you got physically sick?
- What do you think about the following primary care recommendations:
    - Screenings for cholesterol and diabetes?
    - Screenings for cancer?
    - Regular physical exams?
    - Immunizations and vaccinations?
    - Education on your diet, exercise, and lifestyle?
    - Oral exams with the dentist?
- Who do you think is the *most* important person for addressing your physical health problems? Why do you think so?
- How do you prevent your physical illnesses from getting worse over time?
- How do you feel about using primary care in the future?
- Do you have any ideas on how the ACT team can best help you use primary care?

Questions asked to clinicians:

- Could you tell me about the barriers your clients experience in getting primary care?
- (If not touched upon) How do the following factors affect clients' access to primary care?
    - Knowledge of mental illness and substance use disorders among primary care physicians? Discrimination or stigmatization?

    - Psychiatric symptoms?
    - Family relationships?
    - Finances?
- How do you promote primary care among your clients?
- What primary care services would you like to see more of among your clients?
- What is your experience like trying to get clients primary care when they are unwilling to engage with it or go?
- How satisfied are you with your clients' physical health?
- How would you change the ACT team to improve the physical health of your clients?
    - Would you change anything on an individual or interpersonal level?
    - Would you change anything on an institutional or systems level?
- What do you find to be most challenging about improving your clients' physical health?
